# A Continuously Active Antimicrobial Coating Remains Effective After Multiple Contamination Events

**DOI:** 10.1101/2020.09.07.20188607

**Authors:** Luisa A. Ikner, Valerie Beck, Patricia M. Gundy, Charles P. Gerba

## Abstract

Liquid-based disinfection of environmental surfaces is a momentary intervention while the recontamination of these surfaces is continuous. In between disinfection cycles, contaminated surfaces remain a potential source of infection. The use of continuously active antimicrobial surface coatings would reduce the risk of transmission between routine cleaning and liquid disinfection events by serving as an “always-on” approach to reduce pathogen burden. We have recently reported on a surface coating having antiviral properties. Here, the spectrum of activity was broadened assessment efficacy of the coating to withstand multiple contamination events against viruses and pathogenic bacteria.

---

The role of surfaces in disease transmission has been well established (1,2,3). The cleaning and disinfection of environmental surfaces is a critical component in reducing the rate of health care-associated infections (HAI) as demonstrated when effective measures of environmental hygiene are implemented (4–8). The primary method of reducing pathogen exposure from surfaces is through disinfection with a liquid that remains active for only minutes after application. Although liquid-based surface disinfection is effective in the short term, pathogen deposition onto surfaces is a continuous process (9). Low levels of surface contamination have been found to impart an increase in the risk of transmission (5). Many *in vitro* studies of microbial transfer from contaminated surfaces to hands also support these findings (10). Therefore, an intervention that continuously reduces the viability of pathogens deposited onto surfaces between routine cleanings has the potential to disrupt this mode of environmental transmission. The use of antimicrobial surface coatings to serve this purpose have been successfully implemented in the healthcare setting and resulted in reduction of HAI’s (8). We recently reported on an improved formula (SurfaceWise2^TM^) of this continuously active antimicrobial coating technology that reduces levels of infectious human coronavirus 229E (HCoV 229E), an enveloped virus, by 99.99% within 2 hours (Ikner, medrxiv 2020). In the study described herein, this coating technology was found to remain efficacious after multiple contamination events over 8–12 hours against HCoV 229E, as well as Gram-negative and Gram-positive bacteria. This simulates the re-contamination events that a surface would undergo over a typical workday, and at higher microbial load levels.

## Methods

### Test Sample preparation

SurfaceWise2™ (a quaternary ammonium polymer coating) coated surfaces were prepared by Allied BioScience, Inc (Dallas, Texas). Stainless steel coupons (annealed 304) measuring 1” x 1” (used in bacteria studies) and 2” x 2” (used in virus studies) were cleaned with acetone, sterilized by autoclaving, and arranged onto a panel for spraying using a robotic slider equipped with an electrostatic sprayer. Coating coverage was monitored using an XRF (X-Ray Fluorescence Spectroscopy) spectrometer analyzer to ensure uniform application of the coating.

After coating, the coupons were removed from the panel and the coating was cured overnight. Prior to efficacy testing, the coupons were sterilized under UV light for 20 minutes in a biosafety cabinet. Coated stainless steel coupons were held under ambient conditions for 3 days after treatment before use in virus testing and for 2–9 days prior to use in bacterial testing. Non-coated stainless steel carriers were cleaned and sterilized in the same manner for use in the study as control surfaces.

### Preparation of the test virus

Human coronavirus 229E (HCoV 229E, ATCC VR-740), was propagated and assayed using the human lung fibroblast MRC-5 cell line (ATCC CCL-171). Infected cells were subjected to three freeze-thaw cycles to release the virus after cytopathogenic effects (CPE) were observed in the monolayer. The cell lysates were then centrifuged at 1,000 x g for 10 minutes to pellet the cell debris for disposal. Viruses were extracted from the supernatant using polyethylene glycol [9% (w/v), MW 8000] and 0.5 M sodium chloride, with mixing overnight at 4°C. After centrifugation at 10,000 x g for 30 min, the pelleted viruses were resuspended in 0.01 M phosphate buffered saline (PBS; pH 7.4) (Sigma, St. Louis, MO) to approximately 5% of the original virus suspension volume. The virus stocks were then aliquoted and stored at −80°C. HCoV 229E viral stocks were enumerated on MRC-5 cells seeded into 96-well cell culture trays using the TCID_50_ (tissue culture infectious dose at the 50% endpoint) technique as described previously (11). Taking the inverse log of this dilution gives a titer of the virus per ml TCID_50_. The minimum detection for the method described was 3.16 viruses per mL.

### Preparation of bacteria

*Staphylococcus aureus* 6538, *Pseudomonas aeruginosa* 15442, and *Klebsiella aerogenes* 13048 were purchased from American Type Culture Collection (ATCC; Manassas, VA). All strains were re-animated and prepared for storage at –80°C per the vendor’s instructions. Each strain was grown in Nutrient Broth (BD Biosciences; San Jose, CA) at 37°C *(S. aureus* and *P. aeruginosa)* or 30°C *(K. aerogenes)*. Three successive daily transfers were performed for each test organism following incubation at the appropriate temperature, and the final test cultures were incubated for 48 hours. Dilutions were conducted as appropriate for each strain to achieve an approximate inoculation concentration of 5×10^4^ colony-forming units (CFU).

### Test description

The test method was designed to assess the efficacy of a continuously active antimicrobial surface coating when subjected to multiple reinoculations. Between 2 and 6 total inoculations were performed at 2-hour intervals on non-coated control and coated carriers in triplicate. At time zero, all carriers were inoculated the test organism. After 2 hours, one triplicate set each of non-control and coated carriers were neutralized, and all remaining carriers were re-inoculated. After an additional 2 hours (4 hours from time zero), another triplicate set of non-coated control and coated carriers were neutralized, and all remaining carriers were re-inoculated. This pattern continued until the final set of non-coated control and coated carriers completed a 2-hour contact time with the final re-inoculation (Figure 1). For HCoV 229E, efficacy was assessed for 1, 2, 3 and 4 inoculations. For *S. aureus, P. aeruginosa* and *K. aerogenes*, efficacy was assessed for 1, 2, 3, 4, 5 and 6 *(S. aureus* and *P. aeruginosa* only) inoculations.

**Figure 1:**
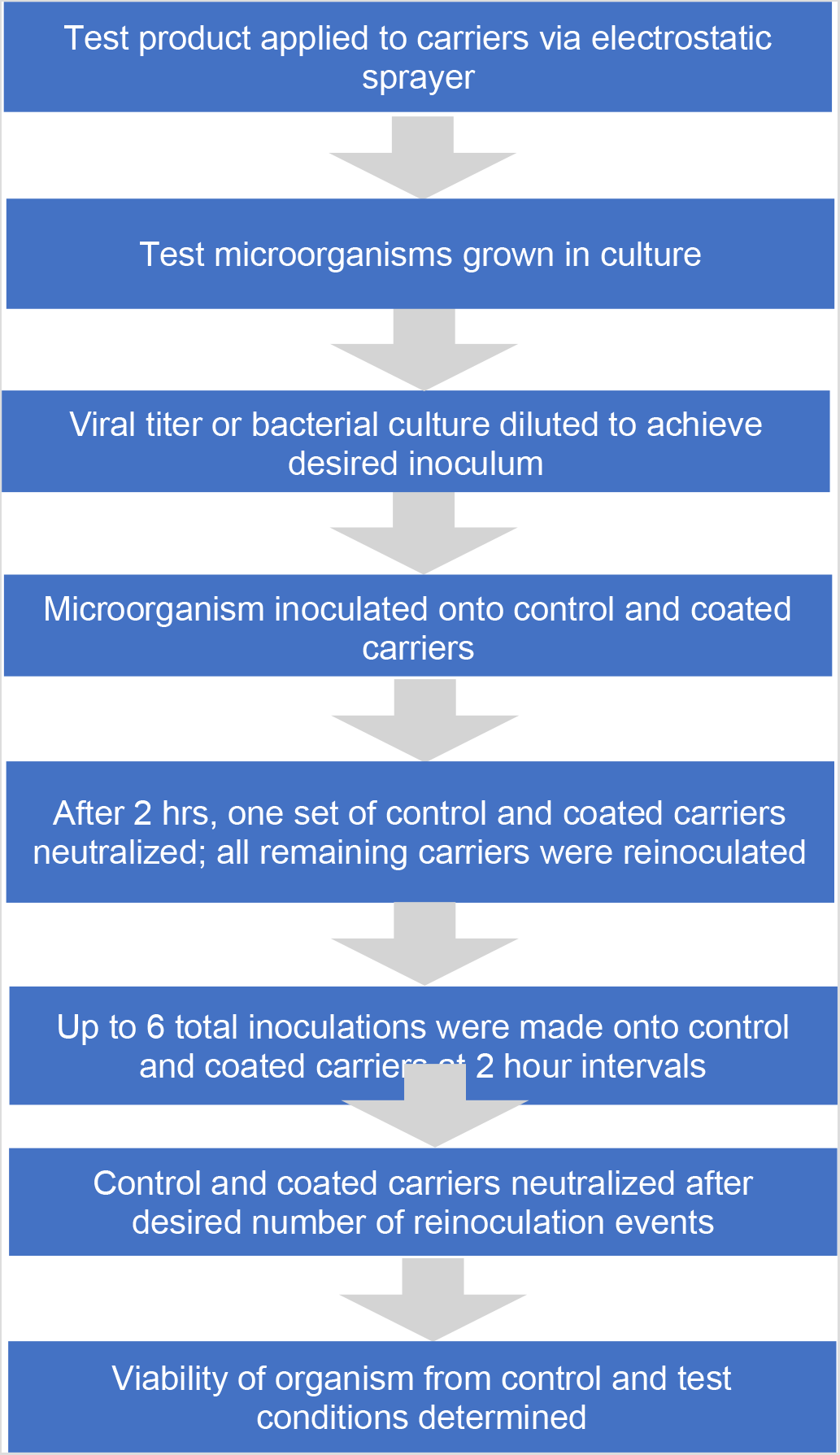
Diagram of the procedure

### Test procedures

#### Virus

A 0.1 mL viral suspension of HCoV 229E with a soil load of 5% FBS (fetal bovine serum) was inoculated and spread over the 2” × 2” surface of control and coated stainless steel carriers, and incubated at room temperature under 45-55% relative humidity. The target initial inoculum for all control and treated carriers was 6.0 log_10_, and 3.5 log_10_ for all subsequent re-inoculations. After each 2-hour contact time, carriers were neutralized by rinsing with 1 mL of Letheen Broth Base (LBB) (Hardy Diagnostics; Santa Martin, CA) with concurrent scraping using a cell scraper tool to further release remaining infectious viruses from the surface. The virus suspension was then immediately passed through a Sephadex G-10 (Sigma; St. Louis, MO) gel filtration column for secondary neutralization and to decrease cytotoxic effects on MRC-5 host cell monolayers. Columns were centrifuged for 5 minutes at 4,000 × g to extract the liquid. Appropriate 1:10 dilutions of neutralized control and test extracts were made using Eagle Minimal Essential Media (MEM) (Mediatech; Manassas, CA), followed by plating onto MRC-5 monolayers prepared in 24-well trays in replicates of four per dilution (0.100 mL per well). The efficacy of the coating was determined by comparing the TCID_50_ levels recovered from the non-coated controls and coated carriers for each group of inoculations. Cytotoxicity and neutralization validations were also performed for confirmation of methodology (data not shown).

#### Bacteria

A 0.01 mL volume of bacterial suspension with a 5% FBS soil load was inoculated and spread over the 1” × 1” surface of non-coated control and coated stainless steel carriers, and incubated at 25°C with 50-55% relative humidity. All inoculations were performed with the same bacterial concentration of approximately 4.7 log_10_. After each 2-hour contact time, the carriers were fully submerged in 20 mL of Dey-Engley (D/E) Neutralizing Broth (Hardy Diagnostics; Santa Martin, CA) and vortexed for 30 seconds to remove remaining viable cells from the surface. Appropriate 1:10 dilutions of neutralized extracts were made using PBS followed by plating onto Tryptic Soy Agar (BD Biosciences; San Jose, CA) using the pour plate method. Plates were incubated at 37°C or 30°C as appropriate for 48 hours before enumeration. Plates with 30–300 colonies were used to determine the CFU per carrier for control and test conditions; the detection limit was 10 CFU per carrier.

## Results

The results shown in Figure 2 demonstrate that the continuously active antimicrobial surface coating reduces viable viral and bacterial levels to below the limit of detection even after multiple contamination events. This translates to a greater than 3-log reduction at all inoculation intervals compared to titers observed for non-coated control surfaces.

**Figure 2.**
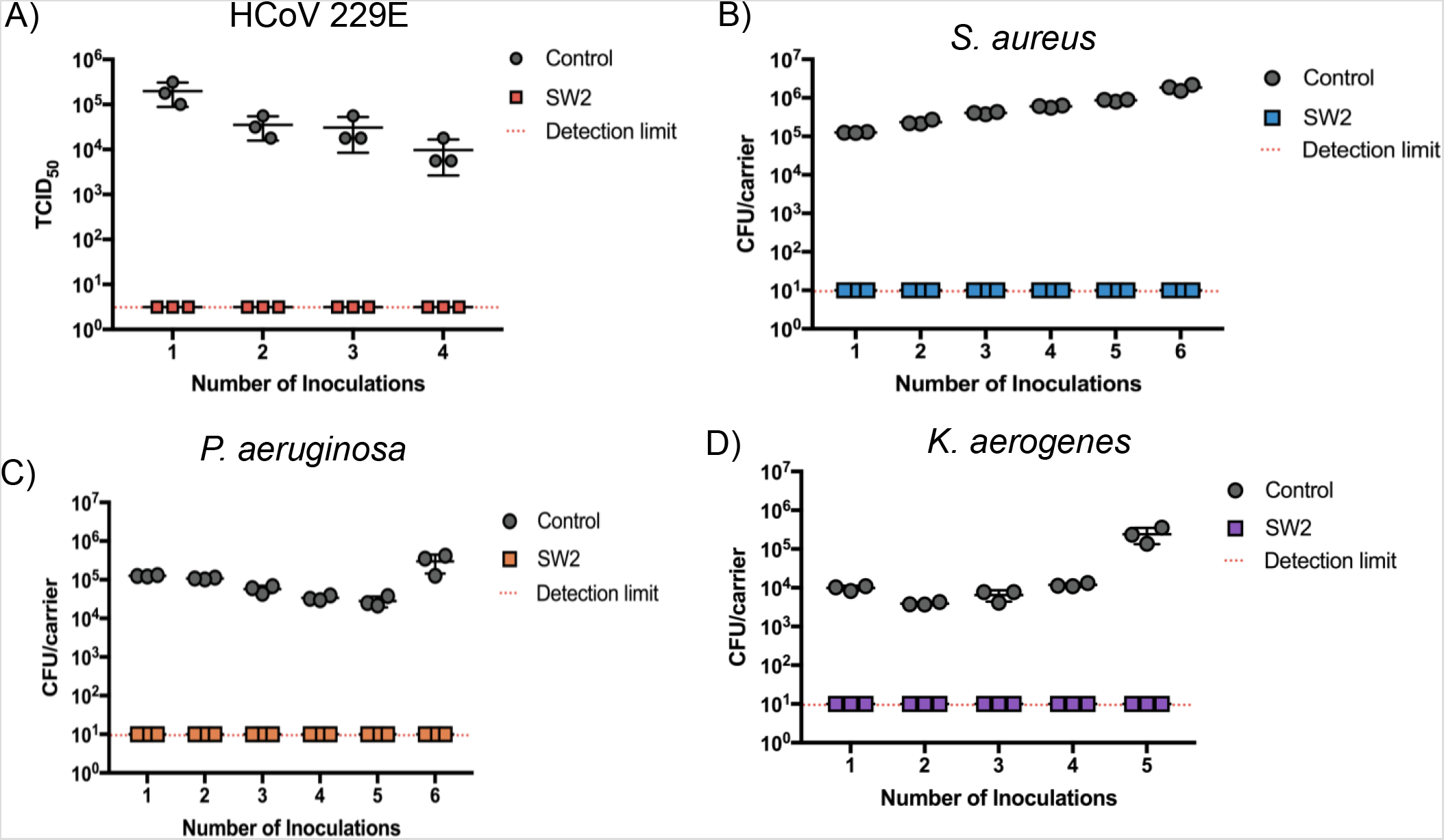
Continuously active antimicrobial surface remains biocidal after multiple contamination events. The efficacy of the antimicrobial surface coating is shown by comparing the amount of virus (A) or bacteria (B-D) recovered from non-coated control (gray circles) and coated (colored squares) stainless steel surfaces.

## Discussion

Antimicrobial coatings that remain active for extended periods of time can be utilized to guard against continual surface contamination events that may occur between routine disinfection cycles. They are intended for use as a supplemental tool to reduce pathogen burden and infection risk from surfaces. Broad spectrum antimicrobial coatings that are efficacious against viruses and bacteria serve as a more robust solution to mitigate infections that might occur due to surface contamination. This study demonstrated that a continuously active disinfectant coating remained efficacious following multiple inoculations of an enveloped virus, and both Gram-positive and Gram-negative bacteria. Such endurance is necessary for practical application in any heavily-trafficked area where large numbers of people come into contact with the same surface multiple times, and cleaning in-between these occasions is neither feasible or practical. In the wake of the COVID-19 pandemic, standard disinfection of surfaces using widely-available disinfectants has been put into practice several times a day to reduce the risk of surface-sourced infections. This approach increases the user’s exposure to these harsh chemicals. Continuously active disinfection technology has the potential to benefit healthcare entities, as well as businesses seeking to re-open to the public following mandated closures, thereby providing a safe and practical alternative to reduce the risk of infection from contaminated surfaces.

## Data Availability

Available to the public upon request

## Reference

1. Boone S, Gerba CP. (2007). Significance of fomites in the spread of respiratory disease and enteric viral disease. Appl Environ Microbiol.(73), 1687–1696.

2. Rutala W, Sickber-Bennet E. (2007). Outbreaks associated with contaminated antiseptics and disinfectants. Antmicrob Agents Chemother (51), 4217–4224.

3. Weber D, Rutala W, Miller M, Huslage K, Sickbert-Bennett E. (2010). Role of hospital surfaces in the transmission of emerging health care-associated pathogens: norovirus, Clostridium difficile, and Acinetobacter species. Am J Infect Control, 38(5 Suppl 1), S25–S33.

4. Boyce JM, Havill NL, Otter JA, McDonald LC, Adams NM, Cooper T, Thompson A, Wiggs L, Killgore G, Tauman A, Noble-Wang J. Impact of hydrogen peroxide vapor room decontamination on Clostridium difficile environmental contamination and transmission in a healthcare setting. Infect Control Hosp Epidemiol. 2008 Aug;29(8): 723–729.

5. Hayden MK, Bonten MJ, Blom DW, Lyle EA, van de Vijver DA, Weinstein RA. Reduction in acquisition of vancomycin-resistant enterococcus after enforcement of routine environmental cleaning measures. Clin Infect Dis. 2006 42: 1552–1560.

6. Dancer SJ, White LF, Lamb J, Girvan EK, Robertson C. Measuring the effect of enhanced cleaning in a UK hospital: a prospective cross-over study. BMC Med. 2009 8; 7: 28. doi: 10.1186/1741-7015-7-28.

7. Carter Y, Barry D. Tackling C difficile with environmental cleaning. Nurs Times. 2011 Sep 13-19; 107: 22–25.

8. Ellingson, K, Pogreba-Brown, K, Gerba, C, & Elliot, S (2019, October 31). Impact of a Novel Antimicrobial Surface Coating on Health Care–Associated Infections and Environmental Bioburden at 2 Urban Hospitals. Clinical Infectious Diseases, 2–1019 ePublication, https://doi.org/10.1093/cid/ciz1077.

9. Kramer A, Schwebke I, Kampf G. How long do nosocomial pathogens persist on inanimate surfaces? A systematic review. BMC Infect Dis 2006; 6: 130.

10. Rusin, P, Maxwell, S, Gerba, C. (2002). Comparative surface-to-hand and fingertip-to-mouth transfer efficiency of gram-positive bacteria, gram-negative bacteria, and phage. J Appl Microbiol, 93: 585–592.

11. Payment, P, and Trudel M. (1993). Methods and Techniques in Virology. New York, NY: Marcel Dekker, Inc.

